# A study of the quality of cardiovascular and diabetes medicines in Malang District, Indonesia, using exposure-based sampling

**DOI:** 10.1101/2022.09.05.22279598

**Authors:** Aksari Dewi, Anushka Patel, Anna Palagyi, Devarsety Praveen, Bachtiar Rifai Pratita Ihsan, Ayuk Lawuningtyas, Diana Lyrawati, Sujarwoto, Asri Maharani, Gindo Tampubolon, Stephen Jan, Elizabeth Pisani

**Affiliations:** The George Institute for Global Health, University of New South Wales, Sydney, Australia; The George Institute for Global Health, University of New South Wales, Hyderabad, India; Department of Pharmacy, Faculty of Medicine, Brawijaya University, Malang, East Java, Indonesia; Department of Public Administration, Brawijaya University, Malang, East Java, Indonesia; Faculty of Biology, Medicine and Health, University of Manchester, United Kingdom; Global Development Institute, University of Manchester, Manchester, United Kingdom

## Abstract

**Background:** The World Health Organization (WHO) has warned that substandard and falsified medicines threaten health, especially in low- and middle-income countries (LMIC). However, the magnitude of that threat for many medicines in different regions is not well described, and high-quality studies remain rare. Recent reviews of studies of cardiovascular and diabetes medicine quality recorded that 15.4 % of cardiovascular and 6.8% of diabetes samples failed at least one quality test. Review authors warn that study quality was mixed. Because they did not record medicine volume, no study reflected the risk posed to patients.

**Methods and Findings:** We investigated the quality of five medicines for cardiovascular disease and diabetes in Malang district, East Java, Indonesia. Our sample frame, based on dispensing volumes by outlet and price category, included sampling from public and private providers and pharmacies, and reflected the potential risk posed to patients. The content of active ingredient was determined by High Performance Liquid Chromatography, and compared with the labelled content. Dissolution testing was also performed.

We collected a total of 204 samples: amlodipine (88); captopril (22); furosemide (21); glibenclamide (21); and simvastatin (52), comprising 83 different brands/products. All were manufactured in Indonesia, and all samples met specifications for both assay and dissolution. None was suspected of being falsified.

**Conclusions:** While we cannot conclude that the prevalence of poor-quality medicines in Malang district is zero, our sampling method, which reflects likely exposure to specific brands and outlets, suggests that the risk to patients is very low; certainly nothing like the rates found in recent reviews of surveys in LMICs. Our study demonstrates the feasibility of sampling medicines based on likely exposure to specific products, and underlines the dangers of extrapolating results across countries.

**What is already known on this topic:** The World Health Organisation suggests that as many as one in 10 medicines in low- and middle-income countries are of poor quality, but studies of the prevalence of substandard and falsified rarely take into account patient exposure.

Medicines for non-communicable diseases and studies from large middle-income countries are under-represented in existing studies.

**What this study adds:** We showed that it is feasible to sample medicines based on patient exposure. Our exposure-based study of cardiovascular and diabetes medicines in Indonesia, a lower-middle income country that is the world’s fourth most populous, found that all met quality standards.

**How this study might affect research, practice or policy:** Adopting exposure-based methods for sampling and/or calculating the prevalence of substandard and falsified medicines would improve our understanding of the potential public health impact of poor-quality products globally.

## Introduction

In 2017, the World Health Organization (WHO) warned that substandard and falsified medicines posed a significant threat to health and to budgets, especially in low- and middle-income countries. The warning, based on data from its newly-strengthened case-reporting system and a review of 100 studies of medicine quality (some unpublished), was summarised in a press release headlined: “1 in 10 medical products in developing countries is substandard or falsified” [1–3].

The WHO noted nine major limitations in its own review, many centring around heterogeneity in definitions, sampling designs and testing. In 2009, scholars proposed Medicine Quality Assessment Reporting Guidelines (MEDQUARG), along with sampling and survey methods [4]. A 2013 review which rated medicine quality studies published between 1948 and 2013 against the MEDQUARG guidelines found that only 15 of 44 meet what the paper’s authors define as minimum standards for research design and reporting (scoring 6 or more on the MEDQUARG checklist) [5]. Standards have improved since the guidelines were published, according to McManus and colleagues, who identified a further 34 studies published between 2013 and 2018; just one of these scored less than 6 [6]. They note, however, that the studies use a variety of sampling methods and quality definitions, complicating the interpretation of results.

MEDQUARG became the basis for methodological guidelines for field surveys of medicine quality published by the World Health Organization (WHO) in 2016 [7]. The guidelines cover various sampling designs (convenience, simple or stratified random sampling, lot quality assurance, and sentinel site monitoring), expressing a preference for random sampling where feasible. More recently, researchers have proposed surveillance methods focused on capturing medicines at highest risk of being substandard [8].

Broadly speaking, these sample designs aim to estimate the prevalence of substandard medicines (which are made by registered pharmaceutical companies in regulated factories but do not meet the quality standards set out in their market authorization paperwork, either because they were poorly made or because they have degraded since manufacture) or of falsified medicines. The latter are made, repackaged, or sold by criminals who seek deliberately to misrepresent the identity, composition, or source of the product [9].

Prevalence of poor-quality medicines is usually expressed as the number of samples failing testing, divided by the number tested, though some designs calculate the proportion of outlets dispensing poor quality medicines [4].

MEDQUARG guidelines suggest reporting information on volumes of sales (potentially allowing the risk of exposure to be calculated), and at least one study has weighted prevalence by sales volume [10]. However, none of the WHO-proposed sampling designs adequately captures the risk posed to patients. For a given level of physical harm caused by a poor-quality medicine, the risk of exposure is determined not only by the prevalence of poor-quality medicines, but also by the likelihood that a patient will consume the type and particular brand of medicine at fault. A small number of brands or outlets may account for a large fraction of patient consumption. In addition, consumption varies by type of medicine and health condition; for example, medicines for chronic conditions are likely to be taken indefinitely, while patients generally only take antimicrobials when experiencing an infection.

### Cardiovascular and diabetes medicines

Medicines for chronic conditions are under-represented among medicine quality studies; just 6.2% of the 48,218 tested medicines included in the WHO review were for non-communicable diseases [2]. A 2019 review identified just five field-based quality surveys including medicines for diabetes, covering 31 countries and totalling 527 samples, of which 6.8% were substandard or falsified[11]. Two of the five surveys used random sampling designs. The medicine most commonly tested in the reviewed studies was metformin; 5.4% of 258 metformin samples collected across four surveys failed at least one quality test. glibenclamide featured in two surveys; 9.2% of 239 samples failed at least one test. A 2021 review of CVD medicine quality studies identified 27 prevalence surveys published between 1996 and 2020. The studies covered 23 active ingredients, in medicines collected in 28 low-or middle-income countries [12]. Overall, 525 out of 3414 samples (15.4%) failed at least one quality test to which they were subjected. However, the authors are careful to note: “we do not state that 15.4% of cardiovascular medicines globally are SF [substandard or falsified]”.

Some 63% of all CVD medicine samples were collected in Africa, many in a study that used stricter criteria for tolerated deviations than permitted by the commonly-used United States Pharmacopeia (USP) standards [13]. Failure rates in Africa were higher than in other regions. Of close to 4,000 samples included in the two reviews, just 212 were collected in Southeast Asia, and only four in Indonesia, the world’s fourth most populous county, where prevalence of hypertension and diabetes among adults aged 45 or more are 52.8% and 13.5% respectively.[14] The four samples, collected between 2009 and 2012, were labelled as a Japanese brand of candesartan; all were judged falsified [15].

### Cardiovascular disease prevention in Indonesia

In an attempt to reduce the burden of CVD in Indonesia, the Ministry of Health has since 2012 supported a prevention and early detection program, including the prescription of medication to prevent cardiovascular events [16]. Members of Indonesia’s nation-wide health insurance system *Jaminan Kesehatan Nasional* or JKN (which at the start of 2022 covered 235.7 million people, around 80% of the population) are entitled to free medication. However, to access it they must follow cumbersome bureaucratic procedures, and medicines are not always available [17,18].

Some Indonesians are thus obliged to buy these medicines, and medicines for other conditions such as diabetes, from pharmacies or elsewhere; others choose to do so for convenience or because they prefer branded medicines which are not provided free. Some vendors do not comply with good pharmaceutical practice, for example in terms of temperature control, or are not regulated by health authorities [19].

Since an auction-based, single-winner procurement platform for JKN medicines known as e-catalogue was introduced in 2014, the volume of medicines procured by the state has risen, and the price paid by Indonesia’s public sector for many essential medicines has fallen dramatically, to levels that producers complain are unsustainably low [20,21]. This, together with a number of medicine falsification scandals in the private sector, raised concerns (expressed in the news media and by professional medical associations) about the quality of the medicines taken by Indonesian patients [22]. Public concern about medicine quality appears at odds with regulatory data. Indonesia’s medicine regulator Badan Pengawasan Obat dan Makanan (BPOM) has been certified by WHO as Maturity Level 3, the second highest level [23]. BPOM is relatively well resourced, with a 2020 budget of US$107 million (72% spent on oversight of medicines and food); over 5,000 staff; and laboratories in every province. Annual post-market surveillance was suspended during the COVID-19 epidemic, but in 2019 BPOM reported 340 of 17,123 sampled medicines were out of specification (1.98%), far below the “1 in 10” intimated by WHO for low and middle income countries [24].

Substandard cardiovascular and diabetes medicines may fail to deliver the correct dosage of active pharmaceutical ingredient (API), thus increasing the risk of cardiovascular events or compromising glucose control, and endangering patients. At a population level, the extent of the threat depends on the number of patients exposed to specific brands of medicine that are poor quality. Because quality may also be affected by handling and storage, the outlet from which medicines are acquired may also influence exposure. However, sampling methods designed to reflect population exposure have not, to our knowledge, been tried in medicine quality surveys, and no studies of the quality of CVD or diabetes medicines in Indonesia exist.

Aiming to fill this gap, we designed a exposure-based study that sampled the five medicines most commonly used by patients at high risk for CVD in eight villages in Malang district, East Java, testing them to ascertain whether they met the quality specifications listed in United States Pharmacopeia and Farmakope Indonesia VI for percent of active ingredients (assay) and for dissolution --a proxy for availability of active ingredients in the body after consumption. Four of these medicines target cardiovascular disease while one was a diabetes medicine, reflecting frequent co-morbidity with the two diseases.

## Methods

We report according to MEDQUARG guidelines. The annotated checklist is available at https://doi.org/10.7910/DVN/EBQYUB, file 01.

### Study setting and background

The study was based around eight villages in Malang district, a district of 2.5 million people in Indonesia’s second most populous province, East Java. The eight villages, which include urban, semi-urban and rural areas, hosted previous research about CVD risk management. [25,26] Researchers screened 99.24% of all adults aged >= 40 in the eight villages in 2018; among the 22,093 people screened, 6,579 adults were identified as at high risk for CVD. For the 2,534 who reported taking any CVD medicine, information was also collected about which medicines they consumed, by API and dosage.

In the study area, patients at high risk for CVD and diabetes may acquire all study medicines for free from public primary health centres, including village-level outreach posts. Most of these medicines are procured through a single national government-run e-catalogue platform and distributed from the District Medicine Warehouse. With rare exceptions (mostly for patented medicines) all are unbranded generics identified by their international non-proprietary name (INN). If the warehouse is out of stock, primary health centres may buy their own INN medicines using capitation funds, a mechanism through which JKN pays public primary health centres and private clinics that accept publicly insured patients a fee per registered participant to deliver preventative services and health care, including medicines.[27] The public hospital provides INN medicines free to JKN-insured patients, paying out of a flat-rate diagnostic-related reimbursement package. Hospitals charge non-insured patients for both INN and branded medicines. They may procure medicines for JKN patients through e-catalogue, independently of the District Medicine Warehouse, or may buy other brands directly from distributors.

Some 50.2% of Malang district residents were JKN members in 2020, well below the national average of 79%. Of those reporting using outpatient services, just 32.7% said they used JKN insurance [28,29]. Most of the remainder sought care from private health care providers --doctors, midwives or nurses. Many doctors provide prescriptions for medicines which patients then buy from pharmacies. A rapid survey of health care providers (see below) indicated that many doctors and midwives also sell prescription medicines directly to patients themselves, although they are not authorized to do so in the study area. A further unauthorised source of medicines are the medicine shops which sell prescription medicines in violation of their over-the-counter-only licenses.

### Sample definition and sample size

Following WHO norms,[7] we defined a single “sample” of medicine as:

- one product (API)
- of one dosage (strength and form)
- of one brand
- from one manufacturer
- and one batch number
- collected at one location, at one time.

Sampling of medicines differs from sampling of individuals, because if good manufacturing and distribution practices are followed, quality should not vary within a batch. Exceptions occur, for example when a genuine batch number is used on a falsified product, or if handling and storage have varied significantly between samples, leading to differential degradation. Broadly speaking, however, a single sample of a medicine should represent the quality of all products of the same API, dose-form and brand, made by the same manufacturer, with the same batch number, sampled in the same location at the same time. A single sample can thus represent the risk of exposure to poor quality medicines for a large proportion of patients.

Our maximum target sample size, determined by budgetary constraints, was of 200 samples, adequate collect at least one sample from all major sources of medicine in the study area (see Figure 1). The Malang District Department of Health gave written permission for the study (070/1102/35.07.103/2020). The study also received ethics approval from the Ethical Committee, Ministry of Research, Technology, and Higher Education, Medical Faculty of Brawijaya University (No.83/EC/KEPK/04/2020) and the Human Research Ethic Committee of University of New South Wales, Sydney (HC200148).). The reflexivity statement in the Supplementary Appendix provides further information about the relationship between institutions. Patients were not directly involved in the design, conduct or reporting of this study.

**Figure 1.**
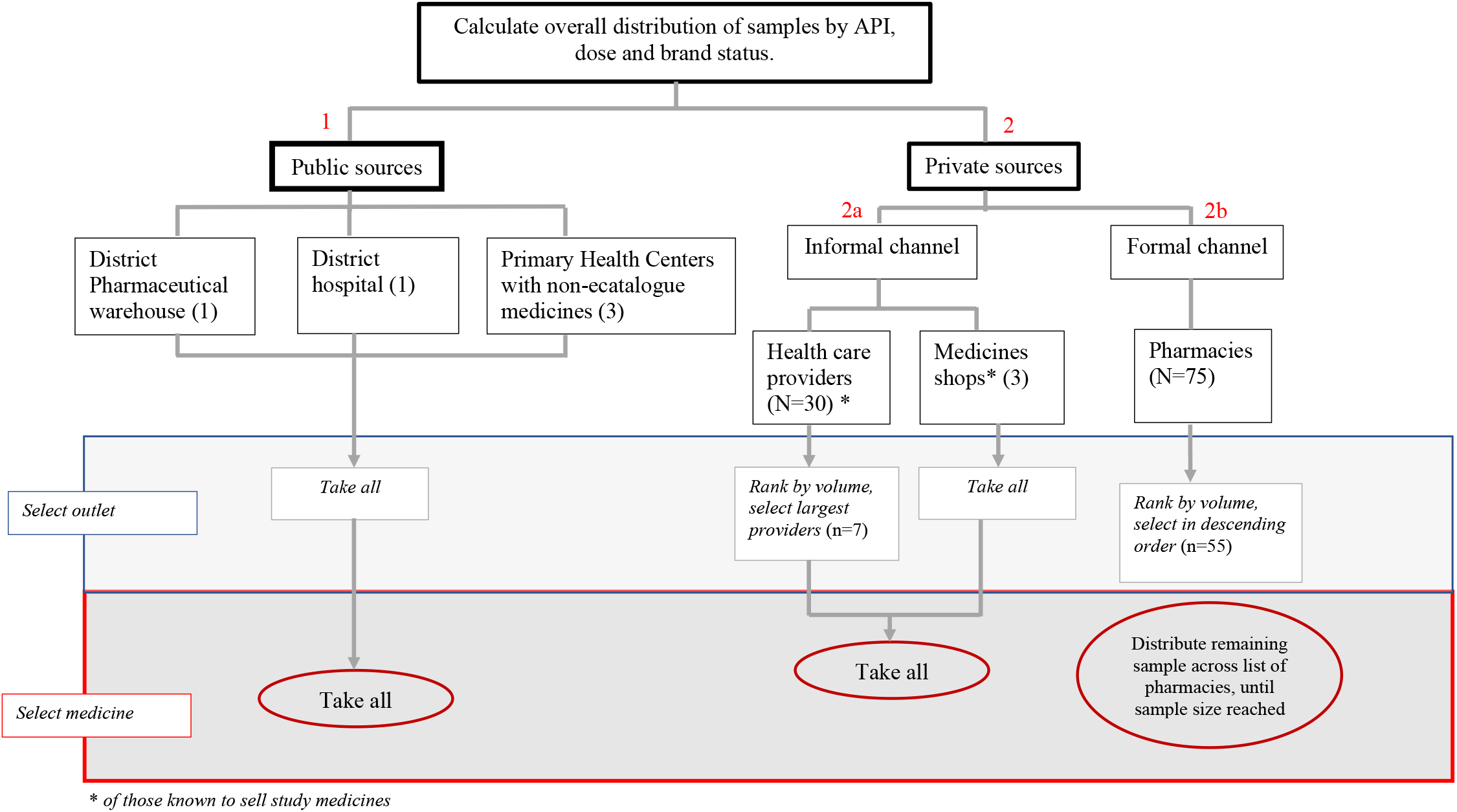
Steps undertaken in construction of sample frame.

### Construction of sample frame: data sources

To construct a sample frame reflecting the likelihood that a patient would take a particular medicine, we collected secondary data from a variety of sources, and also conducted a rapid survey of listed outlets and health care providers. The data, summarised in Table 1, were then triangulated to develop a sample frame reflecting the likely distribution of patients consuming different medicines, by INN status and source of acquisition.

**Table 1:** Data used to inform sample frame.

| Data type | Source | Information provided | Time-frame |
| --- | --- | --- | --- |
| Household survey data | Provided by authors of Reference 25. | % of patients consuming medicine, by API and dose | 2018 |
| Detailed distribution or dispensing data | Secondary data provided by facilities | Volumes distributed or dispensed by API, dose, brand and month: district warehouse; two primary health centres; one private distributor; two pharmacies | April - October 2020 |
| National aggregate sales volume | IQVIA public health | Sales volume by API and dose, by INN status and outlet sector (hospital or retail). | April - October 2020 |
| Listing of pharmacies | District health office, verified by research team | Location of pharmacies and medicine shops in 8 study villages and 4 neighbouring market centres*; private health care providers in 8 study villages | 2019 data received March 2020, verified October 2020 |
| Listing of health care providers and medicine shops | Internet search and public directories, verified by research team | Location of private health care providers and medicine shops in 8 study villages | October – November 2020 |
| Rapid survey of pharmacies and medicine shops | Primary data | Estimate of patients served per day with any study medicine | October – November 2020 |
| Rapid survey of health care providers | Primary data | Estimate of patients served per day with any study medicine, medicines sold, source of medicines | November – December 2020 |

We chose the study medicines based on a 2018 household survey data, in which over 6,500 high-risk patients reported which (if any) medicines they took to control blood pressure or cholesterol.[25] We included all medicines and dosages taken by at least 10% of those reporting medicine use. Because of high levels of co-morbidity, these included one medicine (glibenclamide) to control blood sugar. In order of frequency the medicines were amlodipine, simvastatin, captopril, furosemide and glibenclamide, all in oral tablets. The first three are commonly prescribed in two dosages, the final two in just one, giving a total of eight products (APIs and doses) to be sampled.

With the consent of the management (and where relevant, district health authorities), staff at the District Medicine Warehouse (1/1) and a private medicines distributor (1/40) provided information on volumes of the study medicines distributed each month. Two primary health centres (2/5) and two pharmacies (2/75) provided data on volumes dispensed.

The public health division of health information science company IQVIA provided data on sales volumes of study medicines in Indonesia, disaggregated by INN/branded status. These data are collected on a quarterly basis from a nationally representative panel of >1000 pharmacies, 175 medicine shops, and 250 hospitals in both the private and public sectors.

We obtained listings and contacts of pharmacies, medicine shops and health care providers from sources shown in Table 1. All were contacted in person, and the purpose of the study was explained. From pharmacies, we asked for consent to collect two pieces of information: the estimated number of customers served each day, and the estimated number who were buying medicines for blood pressure, cholesterol, or diabetes. Health care providers gave written consent for brief interviews around medicines provision and procurement. If they reported selling study medicines, we asked for details of dosages, brands, estimated monthly volume and source of medicines, and requested permission to recontact them for possible sampling (see below). Medicine shops were visited to ascertain if they sold study medicines. Most health care providers stated that they sourced their medicines from one of five pharmacies in the two cities nearest to the study area; we added these pharmacies to our sampling list.

### Construction of the sample frame

We constructed a sample frame that reflects the risk that a patient will take any given medicine, by active ingredient, source and brand. For all five study medicines (and eight dosage forms), we triangulated detailed distribution or dispensing data from different sources, dividing volumes by the average number of tablets taken by a patient each month to get an estimated distribution of patients taking each medicine and dose, by branded status. The maximum sample size of 200 was distributed across medicines and dosages to reflect the percent of patients exposed to each medicine and dose.

The overall target was then distributed by sector and outlet type as shown in Figure 1. A detailed explanation of how the same frame was constructed to reflect estimated exposure is provided in the supplementary material, https://doi.org/10.7910/DVN/EBQYUB, File 02.

### Sample collection

Samples were collected between February 3 – May 6 2021. Table 2 summarises sampling methods by facility. Prescriptions were provided by a doctor collaborating with Brawijaya University and were only presented if requested by pharmacists.

**Table 2.** Sampling method by facility.

| Facility | Sampling Method | Acquire by |
| --- | --- | --- |
| District Medicine Warehouse | Overt, with letter from district health authorities | Replacement |
| Primary health centres | Overt, with letter from district health authorities | Replacement |
| District hospital | Overt, with letter from hospital director and formal request letter from university of Brawijaya | Purchase |
| Health care providers | Overt, with letter from district health authorities and formal request letter from university of Brawijaya | Purchase |
| Pharmacies and medicine shops | Mystery shopper, with prescription if requested | Purchase |

For the mystery shopper approach, samples were collected by sample collectors trained using role-play and common vignettes, such as buying medicines for an elderly relative. At each outlet, they requested a single medicine, or a combination consistent with common clinical needs. In order to approximate likely exposure, mystery shoppers did not ask for a specific brand or manufacturer, but rather accepted pharmacists’ suggestions. They did, however, target either branded or unbranded medicines using signalling phrases such as “I’m looking for something affordable” (for INN generics) or mention specificaly that they want to buy generic medicines. For the premium/branded, the mystery shoppers will mention that they want a “patent” product, the term commonly used in Indonesia to signify a premium product.

If the sample frame called for clinically incompatible combinations, or repetitions (for example an INN and a branded version of the same product) from a single outlet, different mystery shoppers were used.

All the study medicines are normally packaged in strips/blisters of 10 tablets. We collected 40 tablets per sample; if 40 tablets were not available, we accepted a minimum of 30 tablets. On exiting the outlet, sample collectors put each sample in a sealable plastic bag marked with a pre-printed barcode. The barcode was scanned and field-related data were entered into a form pre-loaded onto the shoppers’ mobile phones, using open-source KoboCollect software [30]. Further data entry, including product photographs and details of market authorisation holder, manufacturer, registration number and expiry date took place at the end of the day, using a second form linked by the same barcode. The ODK-format data collection forms are available at https://doi.org/10.7910/DVN/EBQYUB Files 03 and 04.

Research team members inspected packaging visually. No reference packaging was available for comparison, so visual inspection, using a magnifying glass as necessary, was limited to checking for anomalies such as mis-spellings, and discrepancies in formatting of batch numbers and expiry dates.

## Sample handling and testing

Samples were stored in a temperature-controlled environment for an average of 21 days, batched and sent (with a temperature logger) for testing to PT Equilab International, an ISO/IEC 17025-certified private laboratory in Jakarta, according to USP 42 NF 37 monograph and using USP reference standards. Methods were validated for all APIs before testing. The full protocols for each molecule are available at https://doi.org/10.7910/DVN/EBQYUB, Files 09-14.

Briefly: laboratory staff inspected tablets visually, noting shape, colour, lettering and other defining characteristics. Chemical analysis was performed for determination of identity, assay (% of labelled active ingredient) and dissolution (% of labelled active ingredient in the tablet dissolved over time). For all APIs, assay testing was by high-performance liquid chromatography, (HPLC -UV; Waters, Aliance 2695with UV Detector 2489 for amlodipine, glibenclamide, furosemide and simvastatin; Waters, Aliance 2695 with Photodiode Array Detector 2996 for captopril)), while dissolution was by Spectrophotometer-UV/VIS (Shimadzu UV-1800) with the exception of glibenclamide, where dissolution was tested by HPLC (Waters, Aliance 2695 with UV Detector 2489).

No testing was performed for uniformity or impurities.

Staff conducting the tests differed from those handling the packaged product, but could see any defining marks on tablets or capsules. Testing took place April – August 2021, an average of 95 days after sample collection.

Results from the certificate of analysis were entered into a database by study staff, using the sample barcode as identifier. Raw dissolution data were added to the database at a later date, delaying stage 2 dissolution. Where necessary, this was undertaken in March 2022.

### Analysis

The KoboCollect field data form, product data form and the laboratory data were merged on barcode number using Stata 17. Stata 17 was also used for reproducible cleaning and coding, and to generate simple descriptive statistics and graphs. The merge and analysis code in Stata format are provided at https://doi.org/10.7910/DVN/EBQYUB, Files 05 and 06.

Table 3 shows the definitions used for compliance with specifications, following USP 42 NF 37 limits, along with the average number of tablets taken by a patient in a month.

**Table 3.** Limits of compliance, United States Pharmacopeia 42 [% of declared content], and average tablets per month used in sample frame calculations.

| API | Assay (%) | Dissolution [Q] (%) | Stage 1 dissolution [Q+5] (%) | Tablets per month |
| --- | --- | --- | --- | --- |
| Amlodipine | 90-110 | 75 | 80 | 30 |
| Captopril | 90-110 | 80 | 85 | 60 |
| Furosemide | 90-110 | 80 | 85 | 30 |
| Glibenclamide | 90-110 | 70 | 75 | 90 |
| Simvastatin | 90-110 | 75 | 80 | 30 |

If any one of six pills included in stage 1 dissolution fell below the Stage 1 threshold of Q+5, we continued to stage 2 testing using additional 6 tablets. The sample was considered out of specification if:

- The assay fell outside the stated limits OR
- Any single tablet fell below the Q threshold -25 in dissolution testing OR
- Any 2 tablets fell below Q treshold -15 in dissolution testing OR
- The average of 12 tablets fell below the Q threshold in stage 2 dissolution testing

## Results

Details of sample frame construction following an exposure-based approach and more detailed information about target sample numbers by medicine, dose and branded status are reported at https://doi.org/10.7910/DVN/EBQYUB, File 02.

Table 4 summarises the number of samples collected from different sources, by INN-branded status In the private sector, we collected a total of 42 unique INN products, and 32 different branded products. Including the public sector, we collected 83 different products (API, dose and brand/market-authorisation holder). All the medicines collected were manufactured in

**Table 4.** Sources of samples collection, by INN or branded status.

| Source | Sector | Outlets | INN samples | Branded samples | Total samples |
| --- | --- | --- | --- | --- | --- |
| District warehouse | Public | 1 | 6 | 0 | 6 (2.9%) |
| District hospital | Public | 1 | 8 | 5 | 13 (6.4%) |
| Primary health centres | Public | 2 | 3 | 0 | 3 (1.5%) |
| Doctor | Unregulated | 4 | 7* | 12 | 19 (9.3%) |
| Midwife | Unregulated | 3 | 3 | 5 | 8 (3.9%) |
| OTC medicine shop | Unregulated | 2 | 6 | 0 | 6 (2.9%) |
| Wholesale pharmacy | Private | 5 | 18 | 8 | 26 (12.7%) |
| Other pharmacy | Private | 55 | 71 | 52 | 123 (60.3%) |
| Total |  | 73 | 122 (59.8%) | 82 (40.2%) | 204 (100%) |

OTC: Over-the-counter
\*Six samples collected, but one sample contained one strip with different package printing, which was tested separately.

Indonesia, and all had valid national market authorisations. Thirty-five samples were packaged in blisters (of which 4 had secondary packaging), the remaining 186 (82.9%) in foil strips.

Mean time to expiry from the date of collection was 674 days in the public sector, 712 days in pharmacies, and 773 days from unregulated sources (private health care providers and medicines shops, who are not technically permitted to sell prescription medicines to patients in Indonesia), with a minimum of 162, 185 and 54 respectively. All samples were tested before expiry.

Retail prices varied by over 100-fold between brands for some medicines, and even the identical product saw up to 10-fold differences in price between retail outlets. Analysis of these data will be reported in detail elsewhere.

### Observations from the field

We found fewer branded generics than expected on the basis of the national market data we used to construct the sample frame (details at https://doi.org/10.7910/DVN/EBQYUB, File 02). Prescriptions are technically required for all study medicines, but sample collectors were instructed to present prescriptions only if requested. None of the 55 retail pharmacies we bought medicines from asked to see a prescription for any medicine.

Daytime temperatures in the study area at the time of data collection ranges between 28 and 30 degrees centigrade, bordering on the unsafe range for storage of medicines. Packaging for all study medicines stipulated that the product should be stored below 30°C. Only 2 of 60 pharmacies (one wholesale and one other) were airconditioned at the time of our visits.

In basic visual inspection, we found a few anomalies, such as strips with two to three tablets in a one-tablet pocket, expiry dates that easily rubbed off, and one medicine with identical batch numbers but with variations in printing techniques for batch number and other information.

### Pharmacopeial testing results

The entire dataset including laboratory results, with brand names masked according to the terms of the ethics approval, is available in xlsx format at https://doi.org/10.7910/DVN/EBQYUB, File 07.

All samples contained the labelled active ingredient. Figure 2 shows the results of assay testing, by API and dosage. Generic INN products are represented by circles, and branded products by diamonds.

**Figure 2:**
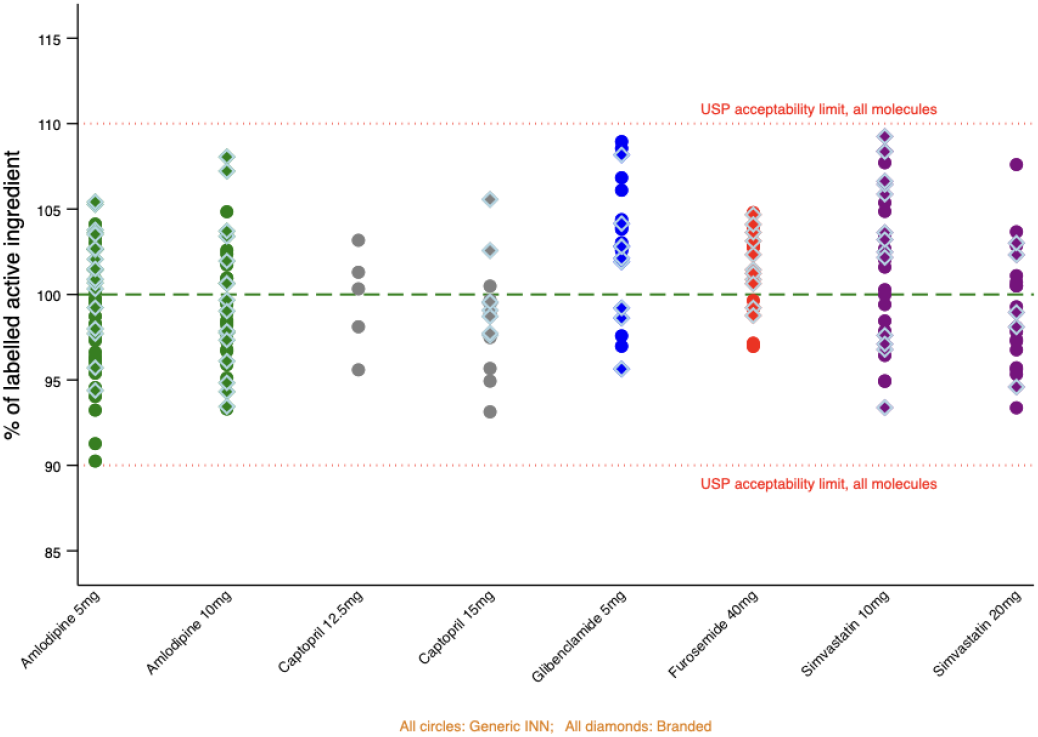
Results of assay testing, by API, dose and INN status.

Assay values ranged from 90.3 to 109.3%, meaning that all samples fell within the USP 42 NF 37 (and Farmakope Indonesia VI) criteria for acceptability which is 90-110%.

Dissolution testing was performed on 203/204 samples. Dissolution parameters differ for different study medicines, as shown in Table 3. While the certificate of analysis showed that average dissolution of the first 6 pills exceeded the required value for all samples, a later review of raw data showed that for 16 samples, not every individual met the overall acceptability threshold plus 5%. Twelve of these samples passed at second dissolution, meaning that a total of 199 samples were considered acceptable by USP 42 NF 37 and Farmakope Indonesia VI standards. Limited remaining tablets meant we were not able to perform Stage 2 dissolution for the remaining four samples. One had a “twin” sample of the same batch which passed at Stage 1 dissolution. Two more were of a single batch of simvastatin which averaged 81.1% at Stage 1 dissolution, well above the acceptability limit of 75%. For the fourth sample, also simvastatin, the average dissolution value for stage one testing was 87.7. Figure 3 shows the final dissolution results by API and dose.

**Figure 3:**
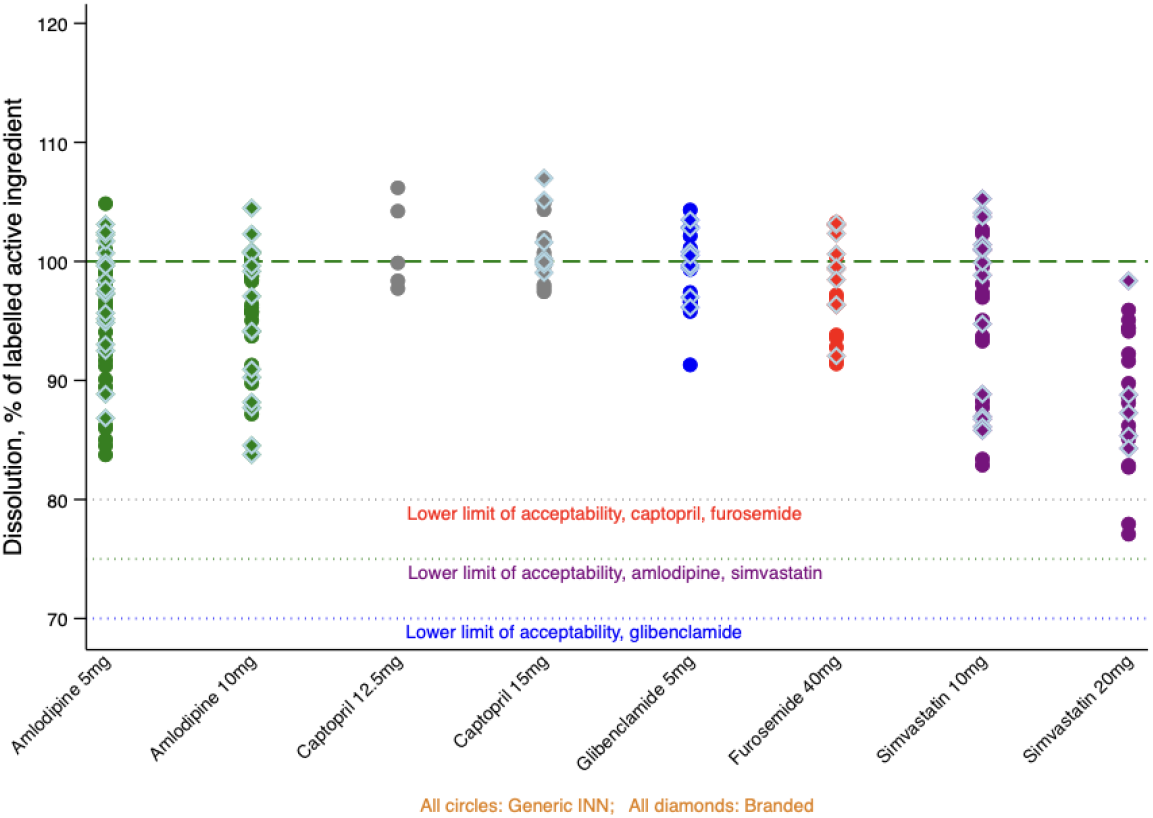
Results of dissolution testing, by API, dose and INN status.

Results were shared with the national and provincial offices of BPOM within a month of the completion of assay and stage 1 dissolution testing.

## Discussion

In our study of 204 samples of 5 common CVD medicines sampled from public, private and informal sources, all were registered; within their expiry dates; all met USP 42 and Farmakope Indonesia VI specifications for assay; and 199 met specifications for dissolution. We were unable to complete dissolution testing for the remaining 5 samples but there were no grounds to expect extreme deviations.

The use of mystery shoppers reduced the likelihood that retail sales staff would selectively provide better quality products, while our take-all approach for medicines provided free in public facilities prevented sampling bias there, despite overt sample collection. However, study limitations mean we cannot conclude that the risk of exposure to poor quality CVD medicines for patients in the study area is zero. We did not test for impurities. Sample collection from doctors and midwives was overt, so although we bought a sample of every variety of study medicine they offered, it is possible that they held back medicines they suspected were of poor quality. However, mystery shoppers also obtained samples of medicines from the pharmacies that doctors and midwives reported buying from, with similar results, suggesting that if bias did exist, it was not considerable.

We did not sample from the internet, or from any of the five private general hospitals in the study area. We do not have data allowing us to estimate the volume of study medicines sold through these channels. However, the additional per-visit consultation fee would likely prevent many patients from choosing to buy medicines for a chronic condition from private hospitals.

Though we checked registration status, we did not have reference packaging, or perform detailed packaging analysis. We are thus unable to rule out falsification, including extension of expiry dates or repackaging of quality INN products to imitate a more expensive brand.

However, there were no out-of-specification products on either assay or dissolution among 83 unique products sampled from 73 outlets, including the district warehouse (which supplies most of the public sector), all of the wholesale pharmacies mentioned as sources of medicine by health-care workers who sell to patients, and 73% of retail pharmacies in the area, including all of the highest volume sellers. We can thus state with confidence that the risk of exposure to poor quality versions of the study medicines is very low in this semi-rural setting in one of Indonesia’s most populous provinces. The situation may differ in other areas of Indonesia. Overall, however, our findings support reports from BPOM’s post-market surveillance which suggest that the overwhelming majority of medicines in the regulated supply chain in Indonesia, including very low-cost unbranded generics in public facilities, meet quality standards.

Our findings differ from those of many previous field surveys in LMICs. The five diabetes prevalence surveys identified by Saraswati et al. in 2019 included 527 samples collected from 31 countries, 382 of them from LMICs. The failure rates in the latter group was 8.6%, compared with 2.1% in high income countries. Within LMICs, failure ranged from 0 (in Chile, CIS, India Pakistan, Thailand and Turkey) to 37.5% in Argentina. However, samples sizes were mostly in single figures. While the failure rate reported for Indonesia was 25%, the 1993 study in question included just four samples from the country, all of glibenclamide. The 27 prevalence surveys for CVD medicines reviewed by Do et al (2021) included 1,889 samples collected in lower-middle income countries, including Indonesia. In this sub-set, prevalence of failure was 16.5%. By country it ranges from 100% failure in Indonesia (4/4 samples) to 0.6% of 521 samples in India (the only other country in the list with a Maturity Level 3 regulator and limited imports of generic medicines) [15,31,32]. In the lower-middle income group, 63.5% of samples were from Africa, with a failure rate of 24.4%. The remainder were from Asia, with a failure rate of 2.9%.

We thus find it difficult to agree with the conclusions of Redfern et al. in their 2019 study of antihypertensive drugs in lower-middle income Nigeria, that “a representative sample from 3 chosen Nigerian states is highly relevant and potentially generalizable across Africa and other developing countries [33].” Indonesia has a large domestic pharmaceutical industry, and all authorised versions of the study medicines are manufactured locally [34]. Currently, the global market for medicines works on a “buyer beware” system, and national regulatory authorities are not responsible for the quality of medicines made for export [35]. Unless countries that rely heavily on imported medicines can police their quality at import, they may thus be exposed to substandard medicines produced elsewhere without adequate regulatory oversight. We speculate that Indonesia’s success in securing the quality of CVD medicines may be in part related to the production-to-market supervision by a single, relatively well-resourced regulator. However, we also note that not all regulations or best practices are observed; we were able to buy all samples without prescriptions, some from sources not permitted to sell these medicines, and most from pharmacies that were not temperature-regulated.

Initial exploration of pricing data indicates that same company may sell a product at very different price points, often producing one or more brands as well as INN versions. This allows for cross-subsidisation across a company’s portfolio, potentially protecting the quality of very low-cost products in the Indonesian market. We plan further investigation of this topic.

Obtaining requisite permissions to collect secondary data for the construction of the exposure-based sample frame, as executed, was time-consuming but feasible. Because no substandard products were found, we were unable to proceed with more detailed estimates of exposure as originally planned. Schiavetti and colleagues, weighting the results of their study of medicines sampled from distributors in the Democratic Republic of Congo by market size, found that those with larger distributions were more likely to be of good quality [10]. We suggest exposure-based sampling could be repeated in settings known to have more poor-quality products, in order to better estimate the true population exposure to substandard and falsified medicines.

## Supporting information

Supplementary_1_MEDQUARG

Supplementary_2_Sampling

## Data Availability

The sample level data are available at https://doi.org/10.7910/DVN/EBQYUB, File 07. In accordance with the terms of the ethics approval, names of individual manufacturers and batch numbers are masked, product registration numbers removed, and outlets grouped by type. Data are provided for reuse, with the expectation that users will cite the dataset and this paper.

https://dataverse.harvard.edu/dataset.xhtml?persistentId=doi:10.7910/DVN/EBQYUB

## Acknowledgements and source of funding

This research was funded by the National Health and Medical Research Council (NHMRC) of Australia under grant number NHMRC APP1149987. We thank Malang District Health Authority, the study participants, and the members of an informal Indonesian medicine supply chain study group whose discussions enriched our understanding of the context. We also thank Slamet Hariono and Elmi Kamilah for their support with data collection. We thank United States Pharmacopeia for providing testing standards at a discounted price for academic research.

