## Supplementary_1_MEDQUARG for "A study of the quality of cardiovascular and diabetes medicines in Malang District, Indonesia, using exposure-based sampling"

### MEDQUARG CHECKLIST

| Item | Section & Topic | Description | Remarks |
|---|---|---|---|
| 1 | Title/abstract/keywords | '- Identify the article as a study of medicine quality (recommended MeSH headings: "medicine quality, substandard, degraded, counterfeit")<br>- Provide an abstract of what was done and what was found, describing the main survey methods and chemical analysis techniques used | Completed |
| 2 | Introduction | - Summarise previous relevant drug quality information and describe the drug regulatory environment<br>- State specific objectives | Completed |
| | Methods | | |
| 3 | Survey details | The timing and location of the survey; when samples collected and when samples analysed. | Completed |
| 4 | Definitions | The definitions of counterfeit, substandard, and degraded medicines used. | Completed |

| | | | |
|---|---|---|---|
| 5 | Outlets | The type, including indices of size (e.g., turnover), of drug outlets sampled. | Reported. We provide volumes (in patient equivalents) for public sector outlets. By the terms of our ethical approval we are not able to indentify individual private sector outlets by name, but provide detailed description of how volumes were considered. |
| 6 | Sampling design | - Sampling design and sample size calculation<br>- Type and number of dosage units purchased/outlet<br>- Definition of sampling frame<br>- Question of interest, assumptions, sampling method(s) (including method of randomisation if random sampling used) | Completed |
| 7 | Samplers | Who carried out the sampling and in what guise? What did the collector say in buying the medicines? | Completed |
| 8 | Statistical methods | Describe the data analysis techniques used. | Completed |
| 9 | Ethical issues | Whether ethical approval was sought and whether the study encountered any ethical issues. | Completed |
| 10 | Packaging | Packaging examination and reference standards. | Reported. We did not have reference packaging for any of the 83 brands included in the sample. |
| 11 | Chemical analysis | Chemical analysis and dissolution testing SOPs and location(s) of laboratory. Description of validation and reference standards used. | Reported in brief. Protocols provided. |

| | | | |
|---|---|---|---|
| 12 | Method validation | Details of laboratory method validation results, including but not limited to: Certificate of analysis for reference standard, within and between run repeatability (RSD% for *n* = 5–8), detection and quantitation limits, accuracy observed for reference samples, linear range for all analytes, sample preparation recovery studies, selectivity. Possibly, validation against a reference method or inter-laboratory study. | All methods validated (reported). No inter-laboratory study was performed. |
| 13 | Blinding | Whether chemistry was performed blinded to packaging and vice versa. | Reported |
| | Results | | |
| 14 | Outlets | The details of the outlets actually sampled, "class" of pharmacy (e.g., public, private for profit, private not for profit, informal, itinerant). | Completed |
| 15 | Missing samples | The reasons why any outlets chosen for sampling did not furnish a sample. Do these outlets differ systematically from those in which samples were obtained? | NA |
| 16 | Packaging and chemistry results | - Packaging and chemistry results and their relationship- Details of products sampled—how many, in what drug classes, countries of origin, batch numbers, manufacture and expiry dates- Results for each analysis—packaging, % AI, dissolution- Additional information could be included in supplementary material | Date of manufactuer is not provided on strip-packaged medicines in Indonesia. All other data are provided as supplementary material at a per sample level. However to comply with the terms of our ethics approval the identity of manufacturers and batch numbers are masked, and outlets are grouped by type. |
| 17 | Category of poor-quality medicine | A clear statement for each medicine sample detected, whether the investigators class it as genuine, counterfeit, substandard, or degraded, with an explanation as to why and whether the medicine was registered with the government in the location(s) sampled. | Completed |

| | | | |
|---|---|---|---|
| 18 | State company and address as given on packaging | If the names of companies and addresses not given, give a reason as to why this information is not provided. | Reported. The terms of our ethics approval require the masking of company names. |
| 19 | Sharing data with MRA | Whether the data shared with the appropriate MRA and IMPACT. | Completed |
| 20 | Dissemination | Description of any non-covert packaging features that would allow others to detect counterfeit medicines. If publication is not possible, consider disseminating via Web-based supplementary material. | NA |
| | Discussion | | |
| 21 | Key results | Summarise key results with reference to study objectives. | Completed |
| 22 | Limitations | Discussion of limitations of study, especially how robust the estimates of prevalence are and how applicable they may be to wider geographical areas. Discuss the direction and extent of any potential bias. | Completed |
| 23 | Interpretation | An interpretation of the results, in conjunction with prior studies, in relation to public health. | Completed |
| 24 | Intervention | Whether interventions are thought appropriate and, if so, what type. | NA |
| | Other Information | | |
| 25 | Conflict of interest | State any potential conflicts of interest. | Completed |
| 26 | Funding | Give the source of funding and role of funders in the study. | Completed |
