## Supplementary_2_Sampling for "A study of the quality of cardiovascular and diabetes medicines in Malang District, Indonesia, using exposure-based sampling"

### Steps taken to construct an exposure-based sample frame for CVD medicines, and details of sampling.

This document accompanies the paper "A study of the quality of cardiovascular and diabetes medicines in Malang District, Indonesia, using exposure-based sampling", by Aksari Dewi and colleagues. It provides a detailed account of the steps taken to develop a sample frame based on the likelihood that a patient will consume a specific brand and dose of a medicine, dispensed by a specific health care provider (public hospital, primary health centre, doctor or midwife or retail outlet) in the study area.

We believe that failure rates derived from a study using this design can be applied to a denominator of the local patient population to estimate the number of patients exposed to poor quality medicines of the types studied.

#### *Data sources*

The sample frame is constructed using primary and secondary data as shown in Table 1 of the accompanying paper.

#### *Construction of the sample frame*

##### Triangulation of data to calculate overall distribution of samples by medicine and dose

For all five study medicines (and eight dosage forms), we triangulated detailed distribution or dispensing data from different sources. Our intention was to estimate the number of patients exposed to poor quality medicines. Because the number of tablets taken per patient varies by medicine, and because one patient may take more than one medicine at a time, we chose to calculate distributions in terms of percent of patients exposed to each medicine, rather than in terms of raw volumes of medicines. We thus divided volumes by the average number of tablets taken by a patient each month (shown in main paper, Table 3) to get an estimated distribution of patients taking each molecule and dose.

Table S1 (provided in .xslx format at https://doi.org/10.7910/DVN/EBQYUB, File 08) shows the data eventually used to construct the sample frame. In the sections below, we break the table down to explain how each row is derived.

| | | | Amlodipine | | Captopril | | Furosemide | Glibenclamide | Simvastatin | |
|---|---|---|---|---|---|---|---|---|---|---|
| | | | 5mg | 10mg | 12.5mg | 25mg | 40mg | 5mg | 10mg | 20mg |
| A1 | Patient survey in 8 study villages | Patients | 1289 | 564 | 170 | 358 | 352 | 247 | 760 | 49 |
| A2 | 2018 data | % | **34.0%** | **14.9%** | **4.5%** | **9.4%** | **9.3%** | **6.5%** | **20.1%** | **1.3%** |
| B1 | District warehouse | Av. patients/month | 2,100 | 745 | 10 | 434 | 173 | 278 | 690 | - |
| B2 | (For all of Kabupaten Malang) | % | 47.4% | 16.8% | 0.2% | 9.8% | 3.9% | 6.3% | 15.6% | 0.0% |
| C1 | IQVIA retail sales | Av. patients/month | 301,931 | 169,402 | 13,988 | 49,086 | 63,079 | 42,507 | 143,514 | 95,493 |
| C2 | (National) | % | 34.3% | 19.3% | 1.6% | 5.6% | 7.2% | 4.8% | 16.3% | 10.9% |
| C3 | % brands in retail outlets that are INN (national data) | | 35.6 | 37.8 | 48.6 | 51.1 | 60.6 | 47.2 | 44.0 | 52.0 |
| D1 | Independent pharmacy, | Av. patients/month | 26 | 35 | 8 | 14 | 7 | 19 | 17 | 10 |
| D2 | Kepanjen (district capital) | % | 19.1% | 25.6% | 5.7% | 10.3% | 4.9% | 14.0% | 12.7% | 7.7% |
| E1 | Chain pharmacy | Av. patients/month | 187 | 187 | 5 | 22 | 49 | 7 | 124 | 105 |
| E2 | Kepanjen (district capital) | % | 27.3% | 27.3% | 0.7% | 3.2% | 7.1% | 1.0% | 18.1% | 15.3% |
| F1 | Local private distributor | Av. patients/month | 6,212 | 6,500 | - | 2,804 | 5,039 | 3,705 | 9,331 | 7,756 |
| F2 | Malang (may sell in other districts) | % | 15.0% | 15.7% | 0.0% | 6.8% | 12.2% | 9.0% | 22.6% | 18.8% |
| G1 | Primary health centre 1 | Av. patients/month | 54 | 11 | 6 | 4 | 3 | 6 | 22 | - |
| G2 | | % | 51.8% | 10.2% | 5.5% | 3.4% | 2.6% | 5.6% | 20.9% | 0.0% |
| H1 | Primary health centre 2 | Av. patients/month | 173 | 17 | 27 | 14 | 3 | 23 | 26 | - |
| H2 | | % | 61.1% | 5.9% | 9.5% | 4.8% | 1.2% | 8.3% | 9.2% | 0.0% |
| X | Unweighted average of Malang based sources, 2020 data | | **36.9%** | **16.9%** | **3.6%** | **6.4%** | **5.3%** | **7.4%** | **16.5%** | **7.0%** |
| Y | Target sample from private sector (N=178), distributed by IQVIA retail proportions (C3) | | **61** | **34** | **3** | **10** | **13** | **9** | **29** | **19** |

**Table S1: Data used to inform a sample frame reflecting risk of exposure to poor quality medicines in Malang district, Indonesia**

**<u>Source A: Household survey of patients in study area, 2018 [1,2]</u>**

**Row A1**: The **number of patients** reporting taking different medicines in the 2018 household survey (see main paper, reference Maharani et al, and Patel et al)

**Row A2**: Calculate **percent distribution of patients per medicine**, of all study medicines.

| | A | B | C | D | E | F | G | H | I | J | K |
|---|---|---|---|---|---|---|---|---|---|---|---|
| 1 | | | | Amlodipine | | Captopril | | Furosemide | Glibenclamide | Simvastatin | |
| 2 | | | | 5mg | 10mg | 12.5mg | 25mg | 40mg | 5mg | 10mg | 20mg |
| 3 | A1 | Patient survey in 8 study villages | Patients | 1289 | 564 | 170 | 358 | 352 | 247 | 760 | 49 |
| 4 | A2 | 2018 data | % | **34.0%** | **14.9%** | **4.5%** | **9.4%** | **9.3%** | **6.5%** | **20.1%** | **1.3%** |

**<u>Source B: Distribution data provided by Malang District Medicine Warehouse.</u>**

This shows distribution to all public outlets in Malang district for March-October 2020.

Row B1: Divide average *monthly distribution* of each API and dosage by number of *tablets taken per patient per month* to get average **number of patients** served by the district warehouse each month.

Row B2: Calculate **percent distribution of patients per medicine**, of all study medicines.

| | | | Amlodipine | | Captopril | | Furosemide | Glibenclamide | Simvastatin | |
|---|---|---|---|---|---|---|---|---|---|---|
| | | | 5mg | 10mg | 12.5mg | 25mg | 40mg | 5mg | 10mg | 20mg |
| B1 | District warehouse | Av. patients/month | 2,100 | 745 | 10 | 434 | 173 | 278 | 690 | - |
| B2 | (For all of Kabupaten Malang) | % | 47.4% | 16.8% | 0.2% | 9.8% | 3.9% | 6.3% | 15.6% | 0.0% |

**<u>Source C: Nationwide retail sales, from data analytics firm IQVIA</u>**

IQVIA collects data quarterly, and provides data on national sales by brand for April to September 2020, in both hospital and retail channels. The volume data provided to us were aggregated by INN status and sales channel; additional information was provided on the % of all marketed products which are branded generics. Data come from a panel of over 1,000 pharmacies selected to be representative at the national level

Row C1: Divide *average monthly retail sales* of each API and dosage by number of *tablets taken per patient per month* to get average **number of patients** buying each medicine at pharmacies per month.

Row C2: Calculate percent distribution of patients per medicine, of all study medicines.

Row C3: Percent of unique products (single API, dose, brand or INN manufacturer) sold in retail outlets that are non-branded INN generics. These data were used to estimate the proportion of branded versus non-branded generics we should aim to sample in pharmacies.

| | | | Amlodipine | | Captopril | | Furosemide | Glibenclamide | Simvastatin | |
|---|---|---|---|---|---|---|---|---|---|---|
| | | | 5mg | 10mg | 12.5mg | 25mg | 40mg | 5mg | 10mg | 20mg |
| C1 | IQVIA retail sales | Av. patients/month | 301,931 | 169,402 | 13,988 | 49,086 | 63,079 | 42,507 | 143,514 | 95,493 |
| C2 | (National) | % | 34.3% | 19.3% | 1.6% | 5.6% | 7.2% | 4.8% | 16.3% | 10.9% |
| C3 | % brands in retail outlets that are INN (national data) | | 35.6 | 37.8 | 48.6 | 51.1 | 60.6 | 47.2 | 44.0 | 52.0 |

**<u>Sources D - H: Detailed sales volume data provided by 2 retail pharmacies and one medicine wholesaler in Malang district</u>**

Rows D1, E1, F1, G1 and H1: Divide *average monthly sales* (D-F) or *dispensed volumes* (G-H) of each API and dosage by number of *tablets taken per patient per month* to get average **number of patients** directly (D, E, G, H) or indirectly (F) served by the outlet each month.

Rows D2, E2, F2, G2 and H2: Calculate **percent distribution of patients per medicine**, of all study medicines.

| | | | Amlodipine | | Captopril | | Furosemide | Glibenclamide | Simvastatin | |
|---|---|---|---|---|---|---|---|---|---|---|
| | | | 5mg | 10mg | 12.5mg | 25mg | 40mg | 5mg | 10mg | 20mg |
| D1 | Independent pharmacy, | Av. patients/month | 26 | 35 | 8 | 14 | 7 | 19 | 17 | 10 |
| D2 | Kepanjen (district capital) | % | 19.1% | 25.6% | 5.7% | 10.3% | 4.9% | 14.0% | 12.7% | 7.7% |
| E1 | Chain pharmacy | Av. patients/month | 187 | 187 | 5 | 22 | 49 | 7 | 124 | 105 |
| E2 | Kepanjen (district capital) | % | 27.3% | 27.3% | 0.7% | 3.2% | 7.1% | 1.0% | 18.1% | 15.3% |
| F1 | Local private distributor | Av. patients/month | 6,212 | 6,500 | - | 2,804 | 5,039 | 3,705 | 9,331 | 7,756 |
| F2 | Malang (may sell in other districts) | % | 15.0% | 15.7% | 0.0% | 6.8% | 12.2% | 9.0% | 22.6% | 18.8% |
| G1 | Primary health centre 1 | Av. patients/month | 54 | 11 | 6 | 4 | 3 | 6 | 22 | - |
| G2 | | % | 51.8% | 10.2% | 5.5% | 3.4% | 2.6% | 5.6% | 20.9% | 0.0% |
| H1 | Primary health centre 2 | Av. patients/month | 173 | 17 | 27 | 14 | 3 | 23 | 26 | - |
| H2 | | % | 61.1% | 5.9% | 9.5% | 4.8% | 1.2% | 8.3% | 9.2% | 0.0% |

#### Triangulation and comparison between sources

We proceeded to compare distributions across the various data sources, using the full table show as Table S1 above, comparing particularly the local data sources (averaged in Row X) and the 2018 survey.

Row X: Calculate the simple average of all data sources derived locally. These include all sources with the exception of IQVIA data, which represent national distributions.

| | | | Amlodipine | | Captopril | | Furosemide | Glibenclamide | Simvastatin | |
|---|---|---|---|---|---|---|---|---|---|---|
| | | | 5mg | 10mg | 12.5mg | 25mg | 40mg | 5mg | 10mg | 20mg |
| A2 | Patient survey, 2018 | % | **34.0%** | **14.9%** | **4.5%** | **9.4%** | **9.3%** | **6.5%** | **20.1%** | **1.3%** |
| B2 | District warehouse | % | 47.4% | 16.8% | 0.2% | 9.8% | 3.9% | 6.3% | 15.6% | 0.0% |
| C2 | IQVIA retail sales | % | 34.3% | 19.3% | 1.6% | 5.6% | 7.2% | 4.8% | 16.3% | 10.9% |
| D2 | Independent pharmacy, | % | 19.1% | 25.6% | 5.7% | 10.3% | 4.9% | 14.0% | 12.7% | 7.7% |
| E2 | Chain pharmacy | % | 27.3% | 27.3% | 0.7% | 3.2% | 7.1% | 1.0% | 18.1% | 15.3% |
| F2 | Local private distributor | % | 15.0% | 15.7% | 0.0% | 6.8% | 12.2% | 9.0% | 22.6% | 18.8% |
| G2 | Primary health centre 1 | % | 51.8% | 10.2% | 5.5% | 3.4% | 2.6% | 5.6% | 20.9% | 0.0% |
| H2 | Primary health centre 2 | % | 61.1% | 5.9% | 9.5% | 4.8% | 1.2% | 8.3% | 9.2% | 0.0% |
| X | Unweighted average of Malang based sources, 2020 data | | **36.9%** | **16.9%** | **3.6%** | **6.4%** | **5.3%** | **7.4%** | **16.5%** | **7.0%** |

The proportionate distribution averaged across the 6 Malang-based sources (row X) was broadly consistent with the consumption data reported in the 2018 household survey (row A2), although the distribution of Simvastatin doses was significantly different. We note that two years after the 2018 survey, the district warehouse had no medicines of this dose in stock. This may suggest that a number of patients prescribed 20mg of Simvastatin were taking 2x10mg because of a localised shortage of 20mg tablets at the time of the 2018 survey. In addition, we noted that the relative distribution of study medicines differed between public and private sectors, with the district warehouse and primary health centres (B2, G2, H2) reporting higher relative dispensing of amlodipine, and the private distributor, pharmacies and IQVIA's national pharmacy sales data (C2-F2) recording more patients supplied with simvastatin.

##### Distribution of total sample size across sectors, medicines, dosages and INN status

Having examined the data, we followed a number of steps in constructing a sample frame. These are summarised in Table S2 (provided in .xslx format at https://doi.org/10.7910/DVN/EBQYUB, File 08), and broken down below.

| | | Amlodipine | | Captopril | | Furosemide | Glibenclamide | Simvastatin | | |
|---|---|---|---|---|---|---|---|---|---|---|
| | 200 | 5mg | 10mg | 12.5mg | 25mg | 40mg | 5mg | 10mg | 20mg | Total |
| Realised 1 | Actual sample public sector | 3 | 4 | 1 | 4 | 4 | 2 | 2 | 2 | **22** |
| Data 1 | Retail sector distribution, IQVIA national | 34.3% | 19.3% | 1.6% | 5.6% | 7.2% | 4.8% | 16.3% | 10.9% | **100%** |
| Target 1 | Target sample, private sector | 61 | 34 | 3 | 10 | 13 | 9 | 29 | 19 | **178** |
| Data 2 | % of brands in private sector INN | 35.6% | 37.8% | 48.6% | 51.1% | 60.6% | 47.2% | 44.0% | 52.0% | |
| Target 2 | Target INN samples, private sector | 22 | 13 | 1 | 5 | 8 | 4 | 13 | 10 | **76** |
| Target 3 | Target branded samples, private sector | 39 | 21 | 1 | 5 | 5 | 5 | 16 | 9 | **102** |
| Realised 2 | Actual INN samples, private sector | 28 | 20 | 4 | 6 | 8 | 10 | 13 | 16 | **105** |
| Realised 3 | Actual branded samples, private sector | 19 | 14 | 0 | 7 | 9 | 9 | 14 | 5 | **77** |
| Realised 4 | Actual sample private sector | 47 | 34 | 4 | 13 | 17 | 19 | 27 | 21 | **182** |
| Realised 5 | **Total final sample** | **50** | **38** | **5** | **17** | **21** | **21** | **29** | **23** | **204** |
| | Number of different brands/producers sampled | 20 | 20 | 2 | 5 | 8 | 6 | 13 | 9 | 83 |

**Table S2: Sample frame: targets and realised samples**

**Step 1: Take-all sample from the public sector**

The public sector has a wider reach than any single retail outlet; the number of patients exposed to any public sector medicine is thus likely to be greater than those buying from a single pharmacy. For this reason, and to minimise any bias that might be introduced by our overt sampling approach, we employed a take-all strategy in the public sector, as follows:

- The district warehouse supplies all public primary health facilities in the district. We aimed to include one sample of every unique version (single API, dose, brand or INN manufacturer) of all study medicines supplied by the district warehouse.
- In case of shortages, primary health facilities may also procure using capitation funds. We planned to collect one sample of every additional unique version (single API, dose, brand or INN manufacturer) of a study medicine from all five health centres in the study area. (We did not re-sample medicines supplied by the district warehouse.)
- The public hospital procures independently, although some of its medicines may also be acquired through the e-catalogue platform, from the same manufacturers as supply the district warehouse. We aimed to collect one sample of every unique version (single API, dose, brand or INN manufacturer) of study medicines offered to public patients by the hospital, other than those that duplicated medicines supplied by the district warehouse. In addition, we planned to collect one sample of each unique branded product sold by the hospital to patients not using public insurance.

The results of this sampling are shown below:

| | | Amlodipine | | Captopril | | Furosemide | Glibenclamide | Simvastatin | | |
|---|---|---|---|---|---|---|---|---|---|---|
| | 200 | 5mg | 10mg | 12.5mg | 25mg | 40mg | 5mg | 10mg | 20mg | Total |
| Realised 1 | Actual sample public sector | 3 | 4 | 1 | 4 | 4 | 2 | 2 | 2 | **22** |

**Step 2: Distribute remaining sample by medicine, dose, and INN status**

We subtracted the 22 samples collected in the public sector from the total target sample of 200. This left 178 samples to be sampled from the private sector. These were distributed by the proportionate distribution of patients served, based on in IQVIA retail data (these were shown in Table S1 Row C2, and are shown below in blue row Data 1), to give the target samples sizes shown in green Row Target 1. We chose to use the IQVIA retail data rather than the 2018 survey data to inform distribution in the private sector because we feared that shortages of simvastatin 20mg in the study area may have led to an overestimate of the proportion of patients taking simvastatin in the 2018 data.

| | | Amlodipine | | Captopril | | Furosemide | Glibenclamide | Simvastatin | | |
|---|---|---|---|---|---|---|---|---|---|---|
| | 200 | 5mg | 10mg | 12.5mg | 25mg | 40mg | 5mg | 10mg | 20mg | Total |
| Realised 1 | Actual sample public sector | 3 | 4 | 1 | 4 | 4 | 2 | 2 | 2 | **22** |
| Data 1 | Retail sector distribution, IQVIA national | 34.3% | 19.3% | 1.6% | 5.6% | 7.2% | 4.8% | 16.3% | 10.9% | **100%** |
| Target 1 | Target sample, private sector | 61 | 34 | 3 | 10 | 13 | 9 | 29 | 19 | **178** |

We then considered the distribution of brands by branded or unbranded (INN) generic status in the private sector, using data shown in Table S1 Row C2, and reproduced below in blue row Data 2. We applied these to the overall target (Target 1), to give the target sample sizes by branded/unbranded status shown in green rows Target 2 and Target 3.

| | | Amlodipine | | Captopril | | Furosemide | Glibenclamide | Simvastatin | | |
|---|---|---|---|---|---|---|---|---|---|---|
| | 200 | 5mg | 10mg | 12.5mg | 25mg | 40mg | 5mg | 10mg | 20mg | Total |
| Target 1 | Target sample, private sector | 61 | 34 | 3 | 10 | 13 | 9 | 29 | 19 | **178** |
| Data 2 | % of brands in private sector INN | 35.6% | 37.8% | 48.6% | 51.1% | 60.6% | 47.2% | 44.0% | 52.0% | |
| Target 2 | Target INN samples, private sector | 22 | 13 | 1 | 5 | 8 | 4 | 13 | 10 | **76** |
| Target 3 | Target branded samples, private sector | 39 | 21 | 1 | 5 | 5 | 5 | 16 | 9 | **102** |

**Step 3: Take-all sample from doctors, nurses and medicine shops.**

Malang district bureau of statistics reports that almost 60% of outpatients go first to private providers, and formative research undertaken during our study planning indicated that many of these doctors and midwives sell medicine to their patients, although they usually stock only a limited variety of medicines and brands.

We used publicly-available information to map doctors and midwives in private practice in the study area, conducted a rapid survey of study medicine sales volumes among those consenting, and ranked them by volume. In order to minimise any bias that might be introduced by our overt sampling approach in this group, we aimed to collect one sample of every unique study medicine (single API, dose, brand or INN manufacturer) sold by all doctors or midwives who reported sales of 300 tablets a month or more.

We mapped and approached 56 health care providers in private practice; 17 did not provide study medicines to patients; 7 others refused interview. We interviewed a total of 32 health care providers (11 doctors, 16 midwives, 5 nurses). Thirty reported selling at least one study medicine; half said they always sold medicines to patients. They reported selling between 30 and 2000 tablets of study medicines a month. We sampled all the study medicines sold by those reporting highest sales volumes (>=300 tablets per month), a total of 19 samples from 4 doctors and 8 samples from 3 midwives.

In our rapid survey of the study villages, we identified three medicine shops not licensed to sell prescription medicines which did in fact sell study medicines. One over-the-counter medicine shop sold all study medicines (with a single product for each) and one other sold one captopril product; we took them all, collecting all 6 samples.

**Step 4: Adjust remainder of sample, and distribute across pharmacies**

We subtracted the samples collected from doctors, midwives and medicine shops (n= 33) from the targets shown for INN or branded generics shown in Table S2 lines Target 2 and Target 3 as appropriate.

We ranked pharmacies by estimated patient numbers for hypertension/cholesterol/diabetes medicines collected in our rapid survey, and distributed the remaining sample size in proportion to that volume, to a maximum of one INN and one branded sample of any medicine/dose combination per outlet. These outlets included "wholesale" pharmacies most frequently reported as supplying health workers (all of them in nearby cities). Twenty-six samples were collected from these wholesale pharmacies, the remainder from retail pharmacies in the study area.

We verified the existence of 75 pharmacies in the study area, and ranked them in order of estimated volumes of CVD customers served per month they provided. To ensure geographic coverage, we first sampled from the highest-volume outlet in each village, drawing several samples of different products, INN and branded, from all but the smallest one. We assigned samples to the remainder of the outlets by patient volume regardless of location, until exhausting the sample size.

We collected 123 samples from 55 pharmacies (73% of those in the study area).

We made a number of adjustments to the targets as sampling progressed. Although national data suggested that branded medicines outnumbered INN products, mystery shoppers found early on that the variety of INN products on offer in the study area was greater than that of branded medicines, perhaps reflecting the semi-rural setting of the study area. Since our aim was to reflect what local residents were likely to use, we adjusted sampling accordingly.

| | | Amlodipine | | Captopril | | Furosemide | Glibenclamide | Simvastatin | | |
|---|---|---|---|---|---|---|---|---|---|---|
| | 200 | 5mg | 10mg | 12.5mg | 25mg | 40mg | 5mg | 10mg | 20mg | Total |
| Target 2 | Target INN samples, private sector | 22 | 13 | 1 | 5 | 8 | 4 | 13 | 10 | **76** |
| Target 3 | Target branded samples, private sector | 39 | 21 | 1 | 5 | 5 | 5 | 16 | 9 | **102** |
| Realised 2 | Actual INN samples, private sector | 28 | 20 | 4 | 6 | 8 | 10 | 13 | 16 | **105** |
| Realised 3 | Actual branded samples, private sector | 19 | 14 | 0 | 7 | 9 | 9 | 14 | 5 | **77** |

In the case of Furosemide and Glibenclamide, the target number of samples was smaller than the 20-sample minimum accepted for testing by the laboratory. We thus increased the total target sample sizes for these two molecules to 20 each, compensating by reducing the sample sizes for Amlodipine, which dominates public sector provision in the study area. We added three additional samples to our expected maximum to accommodate additional Captopril samples.

One four-strip glibenclamide sample contained one strip with the same batch number as the other three but printed in different format. This was split out for separate testing, giving us a total final sample size of 204.

The realised samples from private sources by INN status are shown in orange rows Realised 2 and Realised 3. Table S2 gives the total realised sample size per active ingredient. More information about samples by molecules and source are provided in the main paper, Table 4.
